# Association between cardiometabolic phenotypes with atherogenic index of plasma: a cross-sectional study from the Azar Cohort Study

**DOI:** 10.1101/2024.07.29.24311169

**Authors:** Shireen Soheilifard, Reza Mahdavi, Elnaz Faramarzi

**Author notes:** **Corresponding authors: 1. Reza Mahdavi**: Professor of Biochemistry and Diet Therapy, Faculty of Nutrition and Food Sciences, Tabriz, Iran, P.O. Box: 1567812907, Tabriz, Iran., **2. Elnaz Faramarzi**, Associate Professor, Liver and Gastrointestinal Diseases Research Center of Tabriz University of Medical Sciences, P.O. Box: 1567812907, Tabriz, Iran.

## Abstract

**Background:** This study was conducted to determine the relationship between cardiometabolic phenotypes and atherogenic index of plasma in the Azar cohort population.

**Methods:** This cross-sectional study included 9,515 participants aged 35 to 55, using data from the Azar Cohort Study. Metabolic syndrome was defined based on ATP III criteria. Participants were then classified into four cardiometabolic phenotypes by considering BMI and metabolic syndrome components: metabolically healthy normal weight (MHNW, BMI <25 kg/m²), metabolically unhealthy normal weight (MUHNW, BMI <25 kg/m²), metabolically healthy obese (MHO, BMI ≥25 kg/m²), and metabolically unhealthy obese (MUHO, BMI ≥25 kg/m²). AIP was calculated.

**Result:** Among the subjects, 4,801 (50.5%) were metabolically healthy obese (MHO) and 2,680 (28.2%) were metabolically unhealthy obese (MUHO). High-risk AIP levels (>0.21) differed significantly across cardiometabolic phenotypes, with MUHNW (79.6%) and MUHO (64.6%) showing the highest proportions compared to MHNW (13.5%) and MHO (18.6%). After adjusting for confounders, multinomial logistic regression analysis showed individuals in the third tertile of AIP were 103.46 times more likely to be MUHNW (OR = 103.46, 95% CI: 52.82– 202.64), 55.77 times more likely to be MUHO (OR = 55.77, 95% CI: 45.65–68.12), and 2.22 times more likely to be MHO (OR = 2.22, 95% CI: 1.91–2.64) compared to the MHNW phenotype (p < 0.001 for all).

**Conclusion:** The study demonstrates significant variation in high-risk AIP levels across cardiometabolic phenotypes, emphasizing the need for detailed metabolic health assessments beyond BMI to better predict cardiovascular risk.

## Introduction

Weight is a fundamental measure of health assessment, and its implications are expressed relatively through body mass index (BMI), which adjusts weight for height(1). BMI is widely used to classify people into different weight categories such as underweight, normal weight, overweight, and obese. However, BMI alone does not indicate a person’s metabolic health because it does not distinguish between fat and muscle mass, and individuals with similar BMI but different metabolic health statuses are grouped together(2). For this reason, the grouping of people based on the indicators of metabolic syndrome and BMI has been classified into different phenotypes: Metabolically Unhealthy Obese Normal-weight (MUHNO), Metabolically Healthy Obese (MHO), Metabolically Unhealthy Normal-weight (MUHNW), and Metabolically Healthy Normal-weight (MHNW)(3,4).

Metabolic health is influenced by a wide range of factors, including genetics, lifestyle, diet, and physical activity(5). The complex interplay of these factors makes it challenging to isolate and study specific causes of metabolic health in normal-weight individuals or to understand the transition of obese individuals from being metabolically healthy to metabolically unhealthy. This complexity often requires longitudinal studies and detailed physiological assessments(6). Several studies have reported that BMI or lipid profile assessment alone is not sufficient to diagnose metabolic health; more accurate tools are needed to identify people at risk(7,8).

One of the tools and predictive biomarkers related to metabolic health and cardiovascular risks is the atherogenic index of plasma (AIP)(9). AIP, calculated as the logarithm of the ratio of triglycerides to HDL-C, is associated with the risk of atherosclerosis and cardiovascular disease(10). Previous studies have shown that higher AIP values are associated with increased cardiovascular risk(11,12). However, the relationship between different metabolic phenotypes and AIP has been studied in a very limited way, often yielding inconsistent results or involving small sample sizes, making it difficult to achieve a proper understanding of this relationship(13). Understanding this relationship provides an appropriate platform to guide cardiovascular risk stratification and targeted interventions. **Therefore, to enhance the understanding and accuracy of metabolic health assessments, it is essential and beneficial for public health to conduct studies that correlate cardiometabolic phenotypes with the AIP**. This study was conducted to determine the relationship between different cardiometabolic phenotypes and the atherogenic index of plasma in the Azar cohort population.

## Methods and Materials

### Study design and setting

This cross-sectional study is based on the Azar cohort study, part of the Prospective Epidemiological Research Studies in Iran (PERSIAN)(14). The comprehensive cohort profile describes the initiation of the Azar cohort in 2014, which consisted of three phases: a pilot phase, an enrollment phase, and a 15-year continuous follow-up of participants(15). In the present study, data from the pilot phase and enrollment have been used. The conditions for entering the Azar cohort study were: age 35 to 70 years, at least 9 months of permanent residence in Shabestar city, and having at least one Azeri parent. The exclusion criteria were people with mental or physical disabilities. All participants provided written informed consent, and received detailed information about the study procedure. The inclusion criteria in the present study were people aged 35 to 55 years. The exclusion criteria included pregnant women, people with a history of cancer, and people whose total daily energy intake was more than 4200 kcal or less than 1200 kcal for men, and more than 3500 kcal or less than 1000 kcal for women, as well as those with missing data. Based on these criteria, out of 15,001 participants in the Azar cohort study, a total of 9,515 men and women were included in this study (Figure 1).

**Fig. 1.**
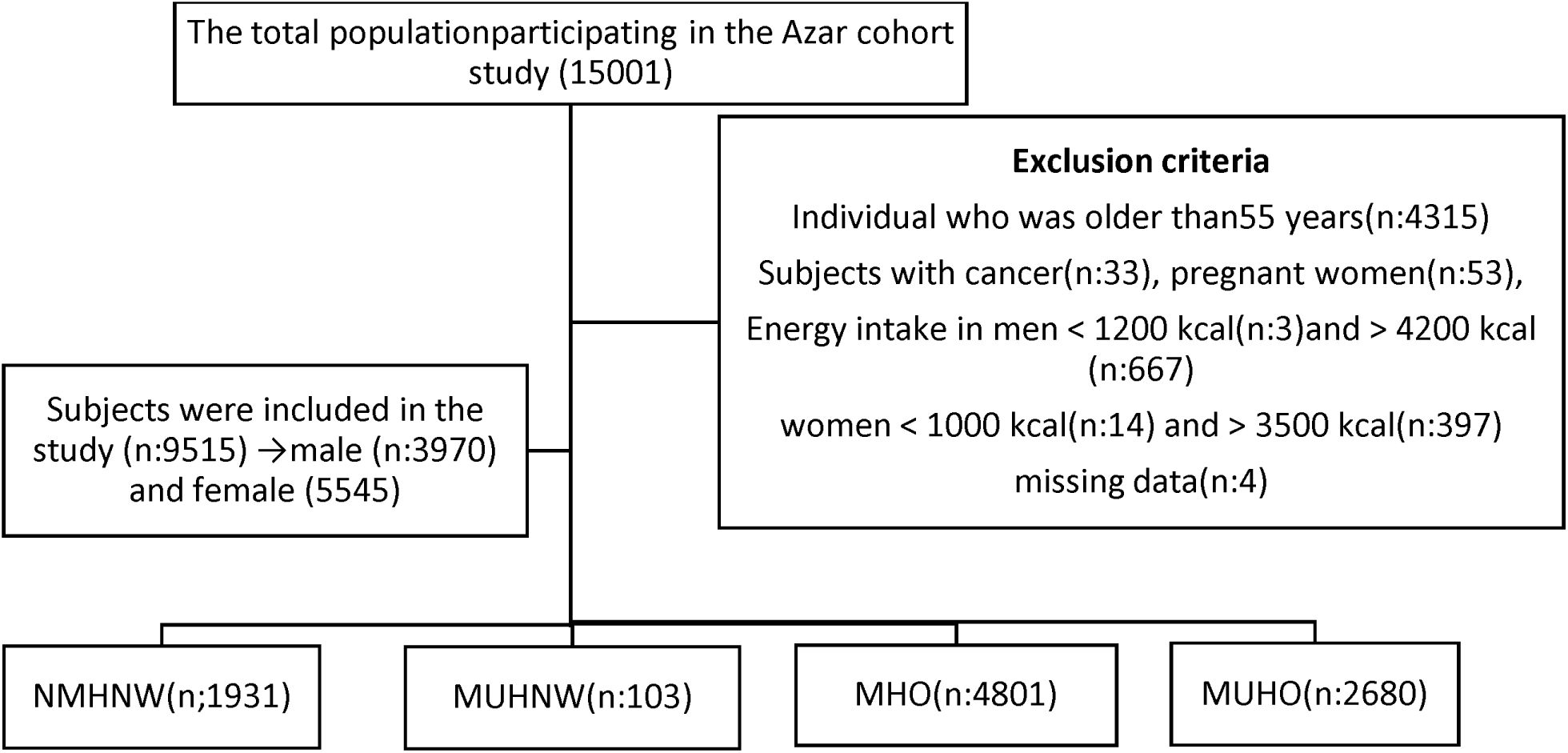
Flow chart of participant selection.

### Ethical statement

This study was approved by the Ethical Board of the Research Council of Tabriz University of Medical Sciences (No.IR.TBZMED.REC.1402.428).

### Data collection

Demographic information including age, gender, marital status, education level, socio-economic status, physical activity, and smoking and alcohol consumption status was collected. The educational status was divided into four groups according to the level and duration of education.

Classification of socio-economic status through the Wealth Score Index (WSI) includes items such as housing conditions (e.g., number of rooms, type of home ownership), available infrastructure (drinking water supply, sanitation facilities), car value, household appliances, electronics, and educational resources. WSI was analyzed using multiple correspondence analysis (MCA). This comprehensive assessment made it possible to classify socio-economic status into five groups from the poorest to the richest.

To evaluate physical activity, participants were divided into three levels: low, moderate, and high, using daily metabolic equivalents (METs). Smoking status was assessed in two categories: non-smoker and smoker. The smoking scale consisted of two options: if respondents reported that they had never smoked, had smoked fewer than 100 cigarettes in their lifetime, or had quit smoking more than a year ago, they were defined as “non-smokers”. If they reported that they were currently smoking or how many cigarettes a day they smoke, they were defined as “smokers”(16). Alcohol consumption was assessed using two options (yes/no).

### Biochemical and clinical measurement

To evaluate biochemical markers, blood samples were collected after a fasting period of 12 to 14 hours. The usual laboratory methods were used to measure levels of total triglycerides (TG), total cholesterol, high-density lipoprotein cholesterol (HDL-C), and low-density lipoprotein cholesterol (LDL-C), using kits from Pars Azmoun, Tehran, Iran. Fasting blood glucose (FBG) was measured through a standard method.

Blood pressure was assessed with a manual sphygmomanometer (Riester) on both arms, with participants seated after a 10-minute rest. The average of the readings was recorded.

#### Anthropometric measurements

The evaluated anthropometric parameters included height, weight, BMI, waist circumference and hip circumference.

Weight was measured using a Seca scale with an accuracy of 0.5 kg. People’s height was measured with meters. The accuracy of the measurements was 0.1 cm. WC was measured with a flexible measuring tape at the level between the lower costal edge and the iliac crest. Hip circumference (HC) was measured around the widest part of the buttocks, with the tape parallel to the floor. Body mass index (BMI) was calculated by dividing weight (kg) by the square of height (meters).

### Definition of different cardiometabolic phenotypes

Metabolic syndrome (MetS) was defined as having at least three criteria specified in NCEP ATP III. These criteria are(17):

- Fasting Blood Glucose (FBG) ≥100 mg/dl or the use of blood glucose-lowering medications
- Serum Triglycerides (TG) ≥150 mg/dl or the use of TG-lowering medications
- High-Density Lipoprotein Cholesterol (HDL-C) ≤40 mg/dl in men and ≤50 mg/dl in women or the use of HDL-C boosting medications
- Systolic Blood Pressure (SBP) ≥130 mm/Hg or Diastolic Blood Pressure (DBP) ≥ 85 mm/Hg or the use of blood pressure-lowering medications
- Waist circumference >102cm in men and >88 in women.

Based on their obesity and metabolic conditions, individuals were categorized into four distinct obesity classifications:

1. metabolically healthy normal weight (MHNW): BMI1<1251kg/m and fewer than three metabolic syndrome components
2. metabolically unhealthy normal weight (MUHNW): BMI1<1251kg/m and at least three metabolic syndrome components
3. metabolically healthy obese (MHO): BMI1≥1251kg/m and fewer than three metabolic syndrome components
4. metabolically unhealthy obese (MUHO): BMI1≥1251kg/m and at least three metabolic syndrome components.

### Calculation Atherogenic Index of Plasma (AIP)

Atherogenic index of plasma was calculated using the formula:

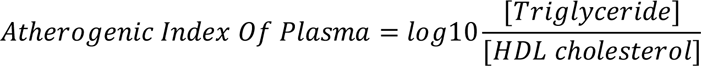

An AIP value <0.11 is associated with a low risk; AIP value from 0.11 to 0.21 indicates an intermediate risk, while AIP >0.21 suggests an increased risk.

### Statistical Analysis

Data analysis was performed using SPSS 14.0 software (SPSS Inc., Chicago, IL, USA). The normality of the data was assessed using Kurtosis and Skewness tests, along with descriptive statistics. Quantitative variables with a normal distribution were reported as mean and standard deviation, while those with a non-normal distribution were reported as median and interquartile range. Qualitative variables were reported as frequency (percentage). One-way analysis of variance (ANOVA) was used to compare quantitative variables between groups and LSD post hoc test for multiple comparisons to determine statistical significance. The Chi-square test and Kruskal-Wallis test were used to compare qualitative variables. Multinomial logistic regression was employed to calculate the odds ratio (OR) and 95% confidence interval (CI) to determine the relationship between different cardiometabolic phenotypes and AIP risk levels (low risk: AIP <0.11, moderate risk: AIP= 0.11- 0.21, and high risk: AIP > 0.21). The model2 was adjusted for age(continues), gender(nominal), WSI (categorical) and physical activity(categorical) as potential confounders, and MHNW and low-risk AIP were considered as reference categories in this analysis. P values <0.05 were considered significant.

## Results

This analysis included 9,515 participants (mean age 45.00 ± 5.89; 58.3% women). Table 1 shows the baseline characteristics of participants stratified cardio metabolic phenotypes. The prevalence of MHNW, MUNW, MHO, and MUHO was 20.3%, 1.1%, 50.5%, and 28.2%, respectively. All baseline variables were statistically significant across cardiometabolic phenotypes(p<0.001). Participants with phenotypes MHO and MHNW were more likely to be younger and have high physical activity levels.

**Table 1:** Baseline characteristics stratified by cardiometabolic phenotypes in the Azar cohort population (n: 9515).

| Variables |  | Total | MHNW<br>n (%) | MUHNW<br>n (%) | MHO<br>n (%) | MUHO<br>n (%) | P |
| --- | --- | --- | --- | --- | --- | --- | --- |
| Gender | Males | 3970(41.7) | 1116(57.8) | 47(45.6) | 1969(41.0) | 838(31.3) | <0.001* |
|  | Females | 5545(58.3) | 816(42.2) | 56(54.4) | 2833(59.0) | 1842(68.7) |  |
| Educations | Illiterate | 772(8.1) | 111(5.8) | 8(7.8) | 338(7.0) | 315(11.8) | <0.001** |
|  | Elementary | 4018(42.3) | 712(36.9) | 40(38.8) | 2023(42.2) | 1243(46.2) |  |
|  | Diploma | 3708(39.0) | 841(43.6) | 42(40.8) | 1913 (39.9) | 912(34.0) |  |
|  | college | 1011(10.6) | 265(13.7) | 13(12.6) | 524 (10.9) | 209(7.8) |  |
| WSI | The lowest | 1919(20.2) | 445(23.0) | 19(18.4) | 878(18.3) | 577(21.5) | <0.001** |
|  | Low | 1346(14.1) | 279(14.4) | 6(5.8) | 669(13.9) | 392(14.6) |  |
|  | Middle | 1895(19.9) | 350(18.1) | 29(28.2) | 1009(21.0) | 507(18.9) |  |
|  | High | 2360(24.8) | 433(22.2) | 27(26.2) | 1255(26.1) | 645(24.1) |  |
|  | The highest | 1995(21.0) | 424(22.0) | 22(21.4) | 990(20.6) | 559(20.9) |  |
| Physical activity<br>(METs) | Low | 2905(30.5) | 526(27.2) | 34(33.0) | 1400(29.2) | 945(35.3) | 0.001** |
|  | Moderate | 3350(35.2) | 589(30.5) | 36(35.0) | 1718(35.8) | 1007(37.6) |  |
|  | High | 3260(34.3) | 816(42.3) | 33(32.0) | 1683(35.1) | 728(27.2) |  |
| Smoking situation | No-Smoker | 8074(84.9) | 1455(75.3) | 81(78.6) | 4155(86.5) | 2383(88.9) | 0.001** |
|  | Smoker | 1441(15.1) | 476(24.7) | 22(21.4) | 646(13.5) | 297(11.1) |  |
| Alcohol consumption | No-Drinker | 9375(98.5) | 1890(97.9) | 102(99.0) | 4745(98.8) | 2638(98.4) | .028** |
|  | Drinker | 140(1.5) | 41(2.1) | 1(1) | 56(1.2) | 42(1.6) |  |

\* p Chi-Square test, \*\* p Kruskal-Wallis test. Abbreviation: MHNW, Metabolically Healthy Normal Weight; MUHN, Metabolically Unhealthy Normal Weight; MHO, Metabolically Healthy Obese, MUHO; Metabolically Unhealthy Obese ;WSI, wealth score index; METs, Metabolic equivalent of task.

Table 2 presents the baseline clinical characteristics and biochemical data for the different cardiometabolic phenotypes. The mean values of AIP and the prevalence of high AIP risk were compared among the four subgroups. The results indicated that both the average AIP and the prevalence of high AIP risk were higher in the MUHNW and MUHO groups compared to the MHNW and MHO groups. Additionally, serum levels of TC, LDL-C, TG,HDL-C, and FBG were higher in the unhealthy metabolic phenotypes than in the healthy metabolic phenotypes. These differences in clinical and biochemical characteristics among the cardiometabolic phenotypes were statistically significant (p<0.001).

**Table 2:** Comparison of anthropometric and biochemical parameters among the studied groups in the Azar cohort population.

| Cardiometabolic<br>Variables |  | Total<br>n:9515 | MHNW<br>n:1931 | MUNHW<br>n:103 | MHO<br>n:4801 | MUHO<br>n:2680 | p |
| --- | --- | --- | --- | --- | --- | --- | --- |
|  |  | Mean±SD | Mean±SD | Mean±SD | Mean±SD | Mean±SD |  |
| Age(year) |  | 45.00±5.89 | 44.05±6.09<sup>a</sup> | 47.49±5.31<sup>b,a,c</sup> | 44.44±5.74<sup>c</sup> | 46.59±5.68<sup>d,a,c</sup> | 0.001* |
| Weight(kg) |  | 76.28±13.55 | 62.13±8.50<sup>a,c,d</sup> | 63.28±7.9<sup>b,c,d</sup> | 78.25±11.24<sup>c,a,d</sup> | 83.43±12.76<sup>d,a,c</sup> | 0.001* |
| Height(cm) |  | 162.81±9.32 | 165.82±9.46<sup>a</sup> | 163.50±8.75<sup>b,a,d</sup> | 162.44±9.18<sup>c,a,d</sup> | 161.27±8.99<sup>d</sup> | 0.001* |
| BMI |  | 28.82±4.89 | 22.53±1.84<sup>a</sup> | 23.59±1.35<sup>b</sup> | 29.65±3.62<sup>c</sup> | 32.07±4.18<sup>d</sup> | 0.001* |
| WC (cm) |  | 93.09±11.08 | 80.21±7.11<sup>a</sup> | 86.62±6.39<sup>b</sup> | 94.10±8.71<sup>c</sup> | 100.78±8.93<sup>d</sup> | 0.001* |
| HC (cm) |  | 104.96±8.81 | 95.74±5.01<sup>a,c,d</sup> | 96.19±4.17<sup>b,c,d</sup> | 106.46±7.34<sup>c</sup> | 109.26±8.60<sup>d</sup> | 0.001* |
| SBP (mmHg) |  | 110.99±15.78 | 104.32±13.55<sup>a</sup> | 117.27±18.44<sup>b,a,c</sup> | 108.83±13.33<sup>c</sup> | 119.43±17.55<sup>d,a,c</sup> | 0.001* |
| DBP (mmHg) |  | 72.98±9.67 | 68.56±8.60<sup>a</sup> | 75.83±10.94<sup>b</sup> | 72.04±8.66<sup>c</sup> | 77.75±10.07<sup>d</sup> | 0.001* |
| FBG (mg/dl) | Mean±SD | 95.98±29.22 | 88.08±17.30 | 122.25±53.54 | 90.10±18.80 | 111.20±41.36 | 0.001*** |
|  | Median (IQR) | 89 (17) | 85.50 (14) | 103.00 (31) | 88.00 (13) | 100.00 (27) |  |
| Total Cholesterol(mg/dl) |  | 190.91±38.80 | 180.38±34.91<sup>a</sup> | 197.73±37.00<sup>b,a</sup> | 190.33±36.89<sup>c,a,d</sup> | 199.27±42.72<sup>d,a,c</sup> | 0.001* |
| LDL-C (mg/dl) |  | 114.79±34.00 | 108.27±31.56<sup>a</sup> | 117.78±33.80<sup>b,a</sup> | 116.00±33.00<sup>c,a</sup> | 117.20±36.79<sup>d,a</sup> | 0.001* |
| TG (mg/dl) | Mean±SD | 149.23±84.09 | 117.46±54.85 | 205.40±75.62 | 130.46±62.11 | 203.65±106.69 | 0.001*** |
|  | Median (IQR) | 123 (74) | 104.00 (36) | 196.00 (94) | 112.00 (42) | 180.00 (97) |  |
| HDL-C(mg/dl) |  | 45.85±10.97 | 48.19±10.97<sup>a, b,d</sup> | 38.30±8.09<sup>b</sup> | 47.78±10.91<sup>c,a,d</sup> | 40.99±8.90<sup>d</sup> | 0.001* |
| AIP | | 0.12±0.25 | 0.0042±0.21<sup>a</sup> | 0.34 ± 0.2<sup>b</sup> | 0.053 ± 0.21<sup>c</sup> | 0.30 ± 0.22<sup>d</sup> | 0.001* |
| AIP, n (%) | low risk | 5051(53.1) | 4919(73.5) | 11(10.7) | 3134(65.3) | 487(18.2) | 0.001** |
|  | moderate risk | 1498(15.7) | 252(13.1) | 10(9.7) | 775(16.1) | 461(17.2) |  |
|  | high risk | 2966(31.2) | 260(13.5) | 82(79.6) | 892(18.6) | 1732(64.6) |  |

\* *p* One Way ANOVA. \*\* *p* Chi-Square test. \*\*\* *p* Kruskal-Wallis. Abbreviation: SD, standard deviation; MHNW, Metabolically Healthy Normal Weight; MUHN, Metabolically Unhealthy Normal Weight; MHO, Metabolically Healthy Obese, MUHO; Metabolically Unhealthy Obese; WC, waist circumference; HC: hip circumference; FBS, fasting blood glucose; HDL, high-density lipoprotein; TG, triglyceride; SBP, systolic blood pressure; DBP, diastolic blood pressure; AIP, Atherogenic index of plasma.
\* *P*-value of the comparison between groups
<sup>a</sup>significant difference between MHNW, MUHNW, MHO and MUHO;
<sup>b</sup>significant difference between MUHNW, MHNW, MHO and MUHO;
<sup>c</sup>significant difference between MHO, MHNW, MUHNW and MUHO;
<sup>d</sup>significant difference between MUHO, MHNW, MUHNW and MHO;
<sup>bac</sup>significant difference between MUHNW, MHNW and MHO;
<sup>dac</sup>significant difference between MUHO, MHNW and MHO;
<sup>acd</sup>significant difference between MHNW, MHO and MUHNW;
<sup>ac</sup>significant difference between MHNW and MHO;
<sup>da</sup>significant difference between MUHNW and MHNW.

Table 3 shows the odds ratio (OR) estimates of AIP risk levels among MUHNW, MHO, and MUHO, with MHNW and low AIP risk as references, using a multinomial logistic regression model. In the unadjusted model, MUHNW and MUHO groups had higher odds of being in the highest AIP level compared to MHNW. Similar results were observed in the adjusted model: the odds of being in the highest third of AIP were 103.46 times higher for MUHNW (OR = 103.46, 95% CI: 52.82–202.64), 55.77 times higher for MUHO (OR = 55.77, 95% CI: 45.65–68.12), and 2.22 times higher for MHO (OR = 2.22, 95% CI: 1.91–2.64) compared to MHNW (p < 0.001 for all).

**Table 3:** The association between cardiometabolic phenotypes and AIP in the Azar cohort population.

| AIP Risk levels | MUHNW |  | MHO |  | MUHO |  |
| --- | --- | --- | --- | --- | --- | --- |
|  | n:103 |  | n:4801 |  | n:2680 |  |
| Model1 | OR (95%CI) | P | OR (95%CI) | P | OR (95%CI) | P |
| Low Risk | Reference |  | Reference |  | Reference |  |
| Intermediate Risk | 5.11 (2.15-12.18) | 0.001 | 1.39(1.19-1.62) | 0.001 | 5.33(4.43-6.41) | 0.001 |
| High Risk | 40.68(21.38-77.39) | 0.001 | 1.55(1.33-1.80) | 0.001 | 19.41(16.44-22.91) | 0.001 |
| *Model2 |  |  |  |  |  |  |
| Low Risk | Reference |  | Reference |  | Reference |  |
| Intermediate Risk | 7.31(3.05-17.50) | 0.001 | 1.64(1.40-1.92) | 0.001 | 7.93(6.48-9.70) | 0.001 |
| High Risk | 103.46(52.82-202.64) | 0.001 | 2.22(1.91-2.64) | 0.001 | 55.77(45.65-68.12) | 0.001 |

*Model 1: Unadjusted model; Model2: Adjusted model for age, gender, physical activity and social economic status.*
*AIP; Atherogenic Index of Plasm, OR, Odd ratio; CI, confidence interval; MUHN, Metabolically Unhealthy Normal Weight; MHO, Metabolically Healthy Obese, MUHO; Metabolically Unhealthy Obese.*

## Discussion

In this large-scale population-based Azar cohort study, our results indicated, the MHO phenotype had the highest frequency at 50.5%, while the MUHNW phenotype had the lowest frequency at 1.1%. Previous research estimated the prevalence of the MHO phenotype to be between 15.1% and 13.3%, and the MUHNW phenotype between 10.5% and 13.3% (18,19). These variations are likely due to differences in genetic factors, age, sex, geographic location, and varying definitions of metabolic health and unhealthiness.

In the current study, the average AIP value for the entire population was 0.12. These results are consistent with those found by Noumegni et al. in Nigeria(20) and Hamzeh et al. in Iran(21). However, they differ from the average AIP values reported by Allahverdiyev et al. in Turkey (0.23), and Tan et al. in Korea(22,23).

Different populations may show different levels of cardiovascular risk factors, potentially due to sample size, environmental, lifestyle, or other background differences(24). For example, in the present study, the prevalence of AIP risk above 0.21 was 31% in the entire population. Considering the large sample size of this research, this represents a significant percentage. These findings are in accordance with the results of studies by Wang et al. in China and Barua et al. in Bangladesh(25,26), but unlike the study by Gol and his et al. in Iran(27). In the current study, the frequency of high-risk levels of AIP was significantly different between cardiometabolic phenotypes. The MUHNW (79.6%) and MUHO (64.6%) groups had a statistically higher proportion of individuals at risk compared to the MHNW (13.5%) and MHO (18.6%) groups. Notably, the MHO group also had a significant number of high-risk participants, indicating a considerable percentage compared to other phenotypes**. We did not find any previous studies that examined the prevalence of different cardiometabolic phenotypes at various AIP risk levels, making our study may be the first to address this aspect.**

The present study showed that all the AIP values in MUHNW (AIP = 0.34) and MUHO (AIP = 0.30) were significantly higher, which corresponded to the average AIP values above 0.21 in these two phenotypes. Similar results were found by Kammar-Garcia et al., who reported that higher AIP was positively and strongly associated with changes in metabolic parameters, and that increased AIP may not be due to increased body mass index alone(28). These values were significantly higher than the average AIP in the MHNW and MHO groups, highlighting the elevated cardiovascular risk associated with metabolic unhealthiness regardless of obesity status(29). These findings align with those reported by Zakerkish et al. and Abdolnezhadian et al., reinforcing the utility of AIP as a reliable marker for identifying individuals at higher risk for cardiovascular diseases(30,31). The significant differences in AIP among the phenotypes underscore its importance in evaluating cardiometabolic health(32,33). In the present study, the frequency of AIP risk levels in the high-risk category was 31%. In contrast, 38% was reported in Wang’s study in China and 57% in Bo et al.’s study in Malaysia(25,34). One of the main reasons for this difference could be the small sample size of these two studies compared to the current one. While sample size can indeed impact the generalizability and variability of study results, it is not the only factor that could account for the observed differences. Other potential reasons might include variations in the demographic characteristics of the study populations, such as lifestyle factors like diet and physical activity, which may influence differences in the prevalence of high-risk AIP.

Another investigation in this study showed a significant relationship between high-risk levels of AIP and cardiometabolic phenotypes regarding the likelihood of developing CVD. After adjusting for potential confounders, individuals in the third tertile of AIP were 103.46 times more likely to be MUHNW, 55.77 times more likely to be MUHO, and 2.22 times more likely to be MHO compared to the MHNW phenotype. The large odds ratio for MUHNW may be due to the small sample size of this phenotype. The results of this study are consistent with the findings of ZakerKish et al., Sadeghi et al. In Iran, Tan et al. in China and several other studies(23,30,35). Previous studies have shown that individuals with higher AIP values are more likely to develop hypertension, dyslipidemia, metabolic syndrome, and cardiovascular disease (CVD)(36–38). Additionally, AIP is crucial for estimating visceral fat content and identifying individuals at high risk for cardiometabolic diseases, such as the MUHO and MUNO phenotypes (39). AIP has proven to be a predictive marker for plasma atherogenicity, closely related to the sizes of HDL, LDL, and VLDL particles, and is a predictor of CVD risk(40,41). AIP is considered the most sensitive marker when compared to three other atherogenic indices: Castelli’s risk index-I (TC/HDL-C), Castelli’s risk index-II (LDL-C/HDL-C), and the atherogenic coefficient (TC-HDL-C/HDL-C(42,43). While elevated triglycerides alone increase the risk of coronary heart disease (CHD), this risk can be mitigated by the cardioprotective effects of HDL cholesterol. Additionally, if other atherogenic risk factors appear normal, AIP can serve as a diagnostic alternative(44).

Our results showed the predictive role of the AIP index in cardiometabolic phenotypes with different BMI. These findings may have important implications for the prevention and control of cardiometabolic diseases. The AIP index could potentially be used as an effective tool to assess cardiovascular disease risk in normal-weight individuals who are sometimes mistakenly thought to be metabolically healthy, as well as in obese phenotypes that are believed to have stable metabolic health.

### Strengths

**The present study is the first to use a large sample size to identify the prevalence and association between the risk levels of the AIP index in metabolically healthy and unhealthy individuals.**

### Limitations

Considering the cross-sectional nature of the study, a causal relationship between different phenotypes and AIP cannot be inferred. The limited number of participants with a normal weight and metabolically unhealthy phenotype compared to other phenotypes makes it difficult to analyze the relationship between this, cardiometabolic phenotype and AIP. Additionally, this study was conducted on a specific population in northwest Iran, which may limit the generalizability of the results to other ethnic or geographical groups.

### Conclusion

The findings of this study elucidate the increased cardiovascular risks associated with the MUHNW phenotype, characterized by a normal BMI but significant metabolic impairment. The high levels of AIP in this group, possibly influenced by lifestyle factors, emphasize the necessity of comprehensive metabolic assessments beyond BMI. On the other hand, the MHO phenotype, while metabolically healthy, still remains at risk for cardiovascular disease. These insights support the need for personalized dietary interventions aimed at improving lipid profiles and reducing atherogenic risk, especially in metabolically unhealthy populations, regardless of BMI. Recognizing and addressing the unique needs of MUHNW and MHO phenotypes can inform public health strategies and clinical interventions aimed at reducing the burden of cardiovascular diseases. In this way, high AIP, in addition to being a biomarker predicting CVD risk, may also indicate the transition from MHO to MUHO. In this regard, a prospective longitudinal study is proposed to investigate the relationship between AIP and the transition of MHO to MUHO over time.

## Declarations

### Consent for Publication

At the beginning of the cohort study, written informed consent for publication was obtained from all participants or their legal guardians.

### Availability of data and materials

The data that support the findings of this study are available from [Vice Chancellor for Research] but restrictions apply to the availability of these data, which were used under license for the current study, and so are not publicly available. Data are however available from the authors upon reasonable request and with permission of [Vice Chancellor for Research]

### Authors’ Contributions

S.S. designed the research. R.M. and E.F. supervised the project, validated the data, and conducted the formal analysis. E.F. curated the data. S.S. analyzed the data and wrote the paper, while R.M. and E.F. reviewed and edited the manuscript. All authors reviewed and approved the final manuscript.

## Acknowledgments

The authors are grateful for the financial support of the liver and gastrointestinal diseases research center, Tabriz University of Medical Sciences. The authors also are deeply indebted to all subjects who participated in this study. We appreciate the contribution by the investigators and the staff of the Azar cohort study. We thank the close collaboration of the Shabestar health center. In addition, we would like to thank the Persian cohort study staff for their technical support. We would like to appreciate the cooperation of the clinical Research Development Unit of Imam Reza General Hospital, Tabriz, Iran in conducting this research.

## Abbreviation

AIP: Atherogenic index of plasma
MHNW: Metabolically healthy normal weight
MUHNW: Metabolically unhealthy normal weight
MHO: Metabolically healthy obese
MUHO: Metabolically unhealthy obese
WSI: Wealth Score Index
MCA: Multiple correspondence analysis
METs: Metabolic equivalents
TG: Triglyceride
HDL-C: High-density lipoprotein cholesterol
LDL-C: Low-density lipoprotein cholesterol
FBG: Fasting blood glucose
WC: Weist circumference
HC: Hip circumference
BMI: Body mass index
SBP: Systolic Blood Pressure
DBP: Diastolic Blood Pressure

## REFERENCES

1. Nuttall FQ. Body Mass Index. Nutr Today. 2015 May;50(3):117–28. 10.1097/NT.0000000000000092

2. Bray GA. Beyond BMI. Nutrients. 2023 May 10;15(10):2254. 10.3390/nu15102254

3. Schulze MB. Metabolic health in normal-weight and obese individuals. Diabetologia. 2019 Apr 19;62(4):558–66. 10.1007/s00125-018-4787-8

4. April-Sanders AK, Rodriguez CJ. Metabolically Healthy Obesity Redefined. JAMA Netw Open. 2021 May 7;4(5):e218860. 10.1001/jamanetworkopen.2021.8860

5. Nedaeinia R, Jafarpour S, Safabakhsh S, Ranjbar M, Poursafa P, Perez P, et al. Lifestyle Genomic interactions in Health and Disease. In 2022. p. 25–74. 10.1007/978-3-030-85357-0_3

6. Mäkinen V-P, Ala-Korpela M. Influence of age and sex on longitudinal metabolic profiles and body weight trajectories in the UK Biobank. Int J Epidemiol [Internet]. 2024 Apr 11;53(3):dyae055. 10.1093/ije/dyae055

7. Shamai L, Lurix E, Shen M, Novaro GM, Szomstein S, Rosenthal R, et al. Association of Body Mass Index and Lipid Profiles: Evaluation of a Broad Spectrum of Body Mass Index Patients Including the Morbidly Obese. Obes Surg [Internet]. 2011;21(1):42–7. 10.1007/s11695-010-0170-7

8. Lafta MA. A comparative study for some atherogenic indices in sera of myocardial infarction, ischemic heart disease patients and control. Journal of Natural Sciences Research. 2014;4(8):96– 103.

9. Fernández-Macías JC, Ochoa-Martínez AC, Varela-Silva JA, Pérez-Maldonado IN. Atherogenic index of plasma: novel predictive biomarker for cardiovascular illnesses. Arch Med Res. 2019;50(5):285–94.

10. Dobiás̆ová M, Frohlich J. The plasma parameter log (TG/HDL-C) as an atherogenic index: correlation with lipoprotein particle size and esterification rate inapob-lipoprotein-depleted plasma (FERHDL). Clin Biochem. 2001 Oct;34(7):583–8. 10.1016/S0009-9120(01)00263-6

11. Dobiasova M. Correspondence to: “Atherogenic index of plasma and the risk of rapid progression of coronary atherosclerosis beyond traditional risk factors.” Atherosclerosis. 2021 Oct;335:148. 10.1016/j.atherosclerosis.2021.09.004

12. Agu PU, Egbugara MN, Ogboi JS, Ajah LO, Nwagha UI, Ugwu EO, et al. Atherogenic Index, Cardiovascular Risk Ratio, and Atherogenic Coefficient as Risk Factors for Cardiovascular Disease in Pre-eclampsia in Southeast Nigeria: A Cross-Sectional Study. Niger J Clin Pract. 2024;27(2):221– 7.

13. Tang M, Zhao Q, Yi K, Wu Y, Xiang Y, Zaid M, et al. Association between Metabolic Phenotypes of Body Fatness and Incident Stroke: A Prospective Cohort Study of Chinese Community Residents. Nutrients. 2022 Dec 9;14(24):5258. 10.3390/nu14245258

14. Poustchi H, Eghtesad S, Kamangar F, Etemadi A, Keshtkar A-A, Hekmatdoost A, et al. Prospective Epidemiological Research Studies in Iran (the PERSIAN Cohort Study): Rationale, Objectives, and Design. Am J Epidemiol. 2018 Apr 1;187(4):647–55. 10.1093/aje/kwx314

15. Farhang S, Faramarzi E, Amini Sani N, Poustchi H, Ostadrahimi A, Alizadeh BZ, et al. Cohort Profile: The AZAR cohort, a health-oriented research model in areas of major environmental change in Central Asia. Int J Epidemiol. 2019 Apr 1;48(2):382–382h. 10.1093/ije/dyy215

16. Naghizadeh S, Faramarzi E, Akbari H, Jafari N, Sarbakhsh P, Mohammadpoorasl A. Prevalence of smoking, alcohol consumption, and drug abuse in Iranian adults: Results of Azar Cohort Study. Health Promot Perspect. 2023 Jul 10;13(2):99–104. 10.34172/hpp.2023.12

17. Alberti KGMM, Eckel RH, Grundy SM, Zimmet PZ, Cleeman JI, Donato KA, et al. Harmonizing the Metabolic Syndrome. Circulation. 2009 Oct 20;120(16):1640–5. 10.1161/CIRCULATIONAHA.109.192644

18. Yang Y, Herting JR, Choi J. Obesity, metabolic abnormality, and health-related quality of life by gender: a cross-sectional study in Korean adults. Quality of Life Research. 2016 Jun 28;25(6):1537–48. 10.1007/s11136-015-1193-2

19. Chen Y, Zhang N, Sun G, Guo X, Yu S, Yang H, et al. Metabolically healthy obesity also has risk for hyperuricemia among Chinese general population: A cross-sectional study. Obes Res Clin Pract. 2016 Sep;10:S84–95. 10.1016/j.orcp.2016.03.008

20. Noumegni SR, Nansseu JR, Bigna JJ, Ama Moor VJ, Kembe Assah F, Dehayem MY, et al. Atherogenic index of plasma and 10-year risk of cardiovascular disease in adult Africans living with HIV infection: A cross-sectional study from Yaoundé, Cameroon. JRSM Cardiovasc Dis. 2017;6:2048004017740478.

21. Hamzeh B, Pasdar Y, Mirzaei N, Faramani RS, Najafi F, Shakiba E, et al. Visceral adiposity index and atherogenic index of plasma as useful predictors of risk of cardiovascular diseases: evidence from a cohort study in Iran. Lipids Health Dis. 2021 Dec 1;20(1):82. 10.1186/s12944-021-01505-w

22. Allahverdiyev A, Koyuncu IMA, Kuru B, Allahverdiyeva A, Ertas FS. The Relationship of Plasma Aterogenity Index and Mean Platelet Volume with the Risk of Development of 1-Year Total Major Adverse Cardiac Event in Patients with Non-ST Elevation Myocardial Infarction. International Journal of Angiology. 2023;32(02):81–7.

23. Tan M, Zhang Y, Jin L, Wang Y, Cui W, Nasifu L, et al. Association between atherogenic index of plasma and prehypertension or hypertension among normoglycemia subjects in a Japan population: a cross-sectional study. Lipids Health Dis. 2023 Jun 29;22(1):87. 10.1186/s12944-023-01853-9

24. BeLue R, Okoror TA, Iwelunmor J, Taylor KD, Degboe AN, Agyemang C, et al. An overview of cardiovascular risk factor burden in sub-Saharan African countries: a socio-cultural perspective. Global Health. 2009;5:1–12.

25. Wang Q, Zheng D, Liu J, Fang L, Li Q. Atherogenic index of plasma is a novel predictor of non-alcoholic fatty liver disease in obese participants: a cross-sectional study. Lipids Health Dis. 2018;17:1–6.

26. Barua L, Faruque M, Banik PC, Ali L. Atherogenic index of plasma and its association with cardiovascular disease risk factors among postmenopausal rural women of Bangladesh. Indian Heart J. 2019 Mar;71(2):155–60. 10.1016/j.ihj.2019.04.012

27. Gol RM, Rafraf M, Jafarabadi MA. Assessment of atherogenic indices and lipid ratios in the apparently healthy women aged 30–55 years. Arterial Hypertension. 2021;25(4):172–7.

28. Kammar-García A, López-Moreno P, Hernández-Hernández ME, Ortíz-Bueno AM, Martínez-Montaño M de LC. Atherogenic index of plasma as a marker of cardiovascular risk factors in Mexicans aged 18 to 22 years. In: Baylor University Medical Center Proceedings. Taylor & Francis; 2021. p. 22–7.

29. Zhu X, Yu L, Zhou H, Ma Q, Zhou X, Lei T, et al. Atherogenic index of plasma is a novel and better biomarker associated with obesity: a population-based cross-sectional study in China. Lipids Health Dis. 2018;17:1–6.

30. Zakerkish M, Hoseinian A, Alipour M, Payami SP. The Association between Cardio-metabolic and hepatic indices and anthropometric measures with metabolically obesity phenotypes: a cross-sectional study from the Hoveyzeh Cohort Study. BMC Endocr Disord. 2023 May 29;23(1):122. 10.1186/s12902-023-01372-9

31. Abolnezhadian F, Hosseini SA, Alipour M, Zakerkish M, Cheraghian B, Ghandil P, et al. Association Metabolic Obesity Phenotypes with Cardiometabolic Index, Atherogenic Index of Plasma and Novel Anthropometric Indices: A Link of FTO-rs9939609 Polymorphism. Vasc Health Risk Manag. 2020 Jun;Volume 16:249–56. 10.2147/VHRM.S251927

32. Dobiasova M, Urbanova Z, Samanek M. Relations between particle size of HDL and LDL lipoproteins and cholesterol esterification rate. Physiol Res. 2005;54(2):159–65.

33. Yang S-H, Du Y, Li X-L, Zhang Y, Li S, Xu R-X, et al. Triglyceride to high-density lipoprotein cholesterol ratio and cardiovascular events in diabetics with coronary artery disease. Am J Med Sci. 2017;354(2):117–24.

34. Bo MS, Cheah WL, Lwin S, Moe Nwe T, Win TT, Aung M. Understanding the Relationship between Atherogenic Index of Plasma and Cardiovascular Disease Risk Factors among Staff of an University in Malaysia. J Nutr Metab. 2018 Jul 4;2018:1–6. 10.1155/2018/7027624

35. Sadeghi M, Heshmat-Ghahdarijani K, Talaei M, Safaei A, Sarrafzadegan N, Roohafza H. The predictive value of atherogenic index of plasma in the prediction of cardiovascular events; a fifteen-year cohort study. Adv Med Sci. 2021 Sep;66(2):418–23. 10.1016/j.advms.2021.09.003

36. Wang L, Chen F, Xiaoqi C, Yujun C, Zijie L. Atherogenic Index of Plasma Is an Independent Risk Factor for Coronary Artery Disease and a Higher SYNTAX Score. Angiology. 2021 Feb 20;72(2):181–6. 10.1177/0003319720949804

37. Ademi Z, Liew D, Zomer E, Gorelik A, Hollingsworth B, Steg PhG, et al. Outcomes and Excess Costs among Patients with Cardiovascular Disease. Heart Lung Circ. 2013 Sep;22(9):724–30. 10.1016/j.hlc.2013.02.002

38. Mirzadeh M, Nikparvar M, Rafati S, Kheirandish M, Azarbad A, Sheybani-Arani M, et al. Atherogenic index of plasma as a predictor of coronary artery disease: a cohort study in south of Iran. The Egyptian Heart Journal. 2024 May 28;76(1):65. 10.1186/s43044-024-00497-z

39. Pinho CPS, Diniz A da S, de Arruda IKG, Leite APDL, Petribú M de MV, Rodrigues IG. Predictive models for estimating visceral fat: The contribution from anthropometric parameters. PLoS One. 2017 Jul 24;12(7):e0178958. 10.1371/journal.pone.0178958

40. Nguyen P, Leray V, Diez M, Serisier S, Bloc’h J Le, Siliart B, et al. Liver lipid metabolism. J Anim Physiol Anim Nutr (Berl). 2008 Jun 28;92(3):272–83. 10.1111/j.1439-0396.2007.00752.x

41. Walther TC, Farese Jr R V. Lipid droplets and cellular lipid metabolism. Annu Rev Biochem. 2012;81:687–714.

42. Tan MH, Johns D, Glazer NB. Pioglitazone Reduces Atherogenic Index of Plasma in Patients with Type 2 Diabetes. Clin Chem. 2004 Jul 1;50(7):1184–8. 10.1373/clinchem.2004.031757

43. Golabi S, Ajloo S, Maghsoudi F, Adelipour M, Naghashpour M. Associations between traditional and non-traditional anthropometric indices and cardiometabolic risk factors among inpatients with type 2 diabetes mellitus: a cross-sectional study. Journal of International Medical Research. 2021;49(10):03000605211049960.

44. Cai X-T, Gao J, Wang M-R, Liu S-S, Hu J-L, Hong J, et al. A J-shaped relationship between the atherogenic index of plasma and new-onset myocardial infarction in hypertensive patients with obstructive sleep apnea: a cohort study. Eur Rev Med Pharmacol Sci. 2022;26(21).

